# Association of BMI and Obesity with Composite poor outcome in COVID-19 adult patients: A Systematic Review and Meta-Analysis

**DOI:** 10.1101/2020.06.28.20142240

**Authors:** Arto Yuwono Soeroto, Nanny Natalia Soetedjo, Aga Purwiga, Prayudi Santoso, Iceu Dimas Kulsum, Hendarsyah Suryadinata, Ferdy Ferdian

## Abstract

**Aim:** This study aimed to evaluate the association between obesity and composite poor outcome in coronavirus disease 2019 (COVID-19) patients.

**Methods:** We conducted a systematic literature search from PubMed and Embase database. We included all original research articles in COVID-19 adult patients and obesity based on classification of Body Mass Index (BMI) and composite poor outcome which consist of mortality, morbidity, admission of Intensive Care Unit (ICU), mechanical ventilation, Acute Respiratory Distress Syndrome (ARDS), and severe COVID-19.

**Results:** Nine studies were included in meta-analysis with 6 studies presented BMI as continuous outcome and 3 studies presented BMI as dichotomous outcome (obese and non-obese). Most studies were conducted in China (55.5%) with remaining studies from French, Germany, and United States (US). COVID-19 patients with composite poor outcome had higher BMI with mean difference 0.55 kg/m^2^ (95% CI 0.07–1.03, P=0.02). BMI ≥30 (obese) was associated with composite poor outcome with odds ratio 1.89 (95% CI 1.06–3.34, P=0.03). Multivariate meta-regression analysis by including three moderators: age, hypertension, and Diabetes Mellitus type 2 (DM type 2) showed the association between obesity and composite poor outcome was affected by age with regression coefficient =-0.06 and P=0.02. Subgroup analysis was not performed due to the limited number of studies for several outcomes.

**Conclusion:** Obesity is a risk factor of composite poor outcome of COVID-19. On the other hand, COVID-19 patients with composite poor outcome have higher BMI. BMI is an important routine procedure that should be assessed in the management of COVID-19 patients and special attention should be given to patients with obesity.

## 1. Introduction

The World Health Organization (WHO) has announced COVID-19 outbreak caused by SARS-CoV-2 as a global pandemic on March 11, 2020. It has rapidly spread across China and many other countries since its first emergence in Wuhan, China on December 2019. Currently, there are 5.404.512 confirmed cases globally with the most cases in the European and American regions and total 343.514 deaths.^1^

COVID-19 exhibits high morbidity and mortality with fatal complications such as ARDS, acute renal injury, shock, and acute cardiac injury.^2, 3^ The elderly, people with underlying diseases, and specific health conditions are more susceptible to infection and prone to serious outcomes.^4^

Obesity results in a dysregulated immune response to respiratory infections.^5^ Obesity also has a great impact on normal lung function. Fat deposits in obesity alter the mechanics of the lungs and chest wall, thus reduce the compliance of the lungs. Many studies also reported excess adiposity is associated with increased production of inflammatory cells and induce airway hyperresponsiveness (AHR). In a patient with ARDS, the work of breathing is increased to meet the body’s oxygen need. This physiological response is complicated by obese state. Thus, obesity may contribute to the increased morbidity associated with obesity in COVID$#x25A1;19 infections.^6^

We aimed to perform a systematic review and meta-analysis to investigate the association between obesity and composite poor outcome in patients with COVID-19.

## 2. Material and Methods

### 2.1 Study Selection and Eligibility Criteria

We included all original research articles in adult COVID-19 patients and body mass index (BMI), obesity and related poor outcomes: mortality, morbidity, admission of ICU, mechanical ventilation, ARDS, and severe COVID-19. Due to the limited number of studies with specific outcome, we generalized all poor outcomes as composite poor outcome. Original research not published in English language, pediatrics subjects (age <17 years old), not available in full text, samples below 30, pediatrics subject, and case reports were excluded from this study.

### 2.2 Literature Search

We conducted a systematic literature search from PubMed and Embase database. We use keywords: (1) “COVID-19” OR “SARS-CoV-2” AND “Characteristics”, (2) “COVID-19” OR “SARS-CoV-2 “AND “Obesity, and (3) “COVID-19” OR “SARS-CoV-2” AND “Body Mass Index” OR “BMI”. We also perform hand searching and exploring the queries through the references cited in some articles in order to include all relevant published articles. We conducted literature research from April 11^th^ and was finalized on May 4^th^ 2020. Duplicate results were removed. The remaining studies were screened for relevance by title and abstract. Further reading and investigation according to inclusion and exclusion was done to search potential relevance studies.

### 2.3 Data Extraction

Data extraction was performed using standardized forms that include generic information (first author, year, place), sample size, study design, age, gender, BMI, and related outcome of interest: mortality, morbidity, admission of ICU mechanical ventilation, ARDS, and severe COVID-19. In addition, we include age and comorbid factors which consist of hypertension, DM type 2, chronic kidney disease (CKD), malignancy, cardiovascular disease (chronic heart failure and coronary artery disease), lung disease (asthma, pulmonary hypertension, chronic obstructive pulmonary disease) for meta-regression analysis. Data extraction was performed independently by two authors (AYS and P).

### 2.4 Statistical Analysis

Stata version 16 was used for data collection and meta-analysis. Effect size for BMI continuous outcome was calculated using Hedges’s g standardized mean difference. Some studies report median and interquartile-range (IQR) instead of mean and standard deviation. In this case, we use approximation formula from Wan et al. for sample size<50 and Cochrane for sample size>50.^7, 8^ Effect size for BMI as dichotomous (<30 kg/m^2^ and ≥ 30 kg/m^2^) outcome were calculated using Mantel-Haenszel formula with random effect model if heterogeneity >75%, otherwise fixed model is preferred and reported as odds ratio (OR). Both BMI as continuous and dichotomous variables reported with its 95% confidence interval. Statistical significance set at ≤0.05 with a two-tailed hypothesis. Meta-regression analysis was performed to examine the impact of moderators: age and comorbid factors which consist of hypertension, and DM type 2, on study effect size both on BMI presented as continuous and dichotomous outcome. Other comorbid factors were not conducted for meta-regression due to inappropriate data.

## 3. Results

### 3.1 Study Selection

Initial search yield 657 records from an electronic database and 26 records from hand-searching, respectively. 159 records identified as duplicate. 447 records were excluded after screening the title or abstract. After evaluating and assessing 77 potential studies, 68 studies were removed due one of the following: not available in English language, no outcome of interest, and not original research article. Nine Studies were included in this meta-analysis with 6 studies presented BMI as continuous outcome and 3 studies presented BMI as dichotomous outcome (<30 kg/m^2^ and ≥ 30 kg/m^2^). The selection process is shown in Figure 1.

**Figure 1.**
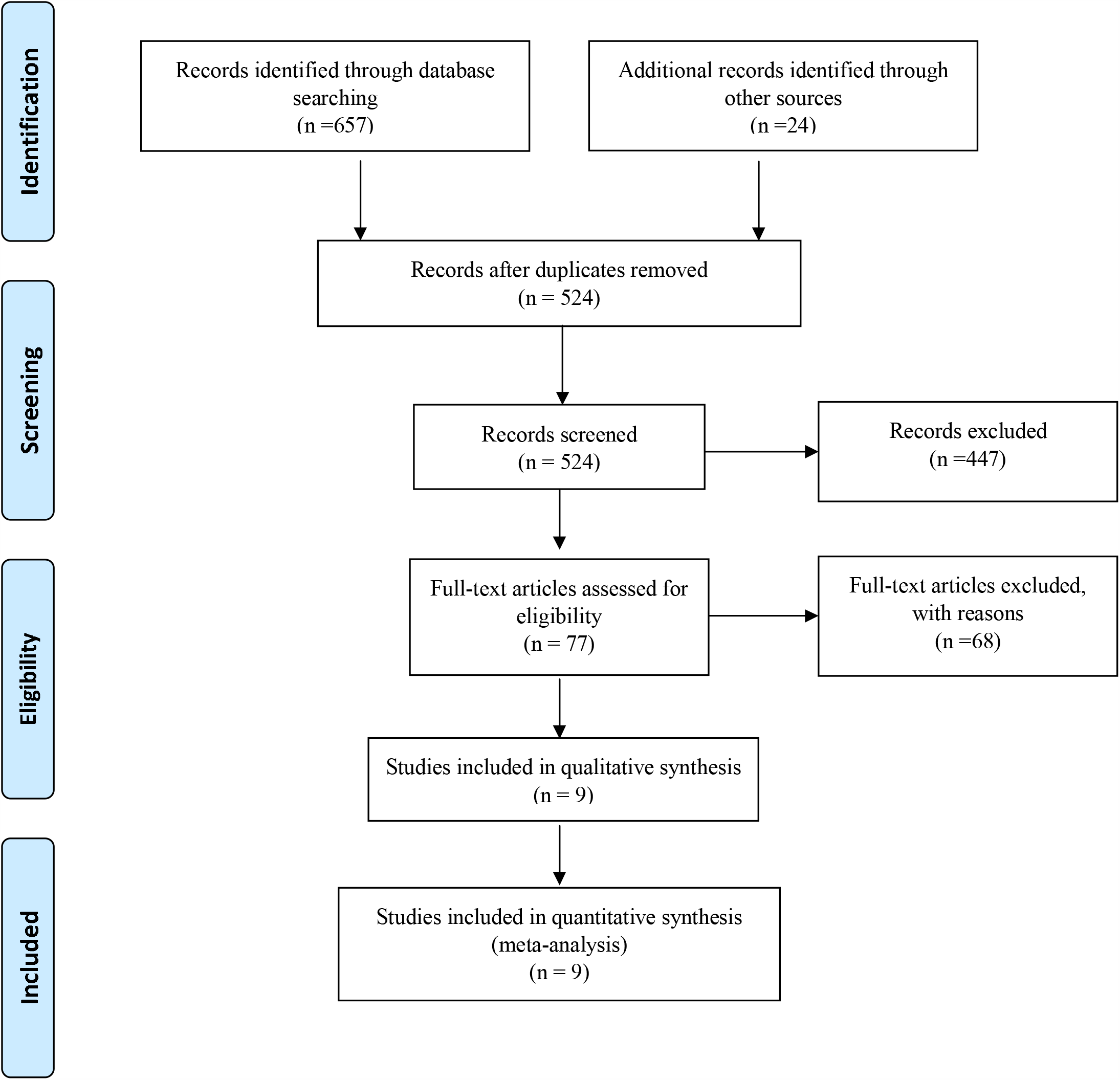
PRISMA Flowchart

### 3.2 Study Characteristics

The basic characteristics of the study are show in Table 1. Most studies were conducted in China (55.5%) with remaining studies from French, Germany, and the US. Severe Covid-19 is the most frequent outcome of interest in this meta-analysis study (44.4%), followed by ICU admission (22.2%). There were a total of 1817 patients from 9 studies.

**Table 1.** Characteristics of Included Studies

| Authors | Study Design | Setting | Samples | Age<br>(Mean/Median) | Male(%) | DM(%) | Hypertension<br>(%) | Measured<br>Outcome |
| --- | --- | --- | --- | --- | --- | --- | --- | --- |
| Cai Q, 2020<sup>9</sup> | Observational<br>prospective | China | 298 (58<br>vs. 240) | 47.5 (62.5 vs.<br>41) | 48.7 (67.2<br>vs. 44.2) | 6 (13.7<br>vs. 4) | 9 (37.9 vs.<br>10.4) | Severe Covid-19 |
| Cao J, 2020<sup>10</sup> | Observational<br>retrospective | China | 102 (17<br>vs. 85) | 54 (72 vs. 53) | 52 (76.5<br>vs. 47) | 10.7<br>(35.2 vs.<br>5.8) | 27.4 (64.7 vs.<br>20) | Mortality |
| Chen Q,<br>2020<sup>11</sup> | Observational<br>retrospective | China | 145 (43<br>vs. 102) | 47.5 (52.8 vs.<br>45.3) | 54.5 (53.5<br>vs. 54.9) | 9.6<br>(15.5 vs.<br>6.8) | 15.1 (20.9 vs.<br>12.7) | Severe Covid-19 |
| Li X, 2020<sup>12</sup> | Observational<br>retrospective | China | 548<br>(269 vs.<br>279) | 60 (65 vs. 56) | 50.9 (56.9<br>vs. 45.2) | 15.1<br>(19.3 vs.<br>11.1) | 30.2 (38.6 vs.<br>59.1) | Severe Covid-19 |
| Simonnet A,<br>2020<sup>13</sup> | Observational<br>retrospective | French | 124 (85<br>vs. 39) | 60 (60 vs. 60) | 73 (75 vs.<br>67) | 22.5 (27<br>vs. 12.8) | 49 (56 vs. 32) | Mechanical<br>Ventilation |
| Kalligeros M,<br>2020<sup>14</sup> | Observational<br>retrospective | US | 103 (44<br>vs. 59) | 60 (61.5 vs. 57) | 61.6 (65.9<br>vs. 57.6) | 36.8<br>(47.7 vs.<br>28.8) | 64.0 (70.4 vs.<br>59.3) | ICU<br>Admission |
| Hu L, 2020<sup>15</sup> | Observational<br>retrospective | China | 323<br>(172 vs.<br>151) | 61 (67 vs. 56) | 51.4 (52.9<br>vs. 49.7) | 14.5<br>(19.1 vs.<br>9.2) | 32.5 (38.3 vs.<br>25.8) | Severe Covid-19 |
| Dreher M,<br>2020<sup>16</sup> | Observational<br>retrospective | Germany | 50 (24<br>vs. 26) | 64.5 (61.4 vs.<br>67.6) | 66 (62 vs.<br>69) | 58 (62.5<br>vs. 53.8) | 70 (67 vs. 73) | ARDS |
| Stavros G,<br>2020<sup>17</sup> | Observational<br>retrospective | US | 124 (60<br>vs. 64) | 64.5 (61.4 vs.<br>67.6) | 61.2 (68.3<br>vs. 54) | 29.8 (30<br>vs 29.6) | 58 (50. vs.<br>64.7) | ICU<br>Admission |

Data compared between outcome (+) and outcome (-) group. ICU: Intensive Care Unit; ARDS: Acute Respiratory Distress Syndrome, US: United States

### 3.3 Body mass index (BMI) and composite poor outcome

Meta-analysis conducted from six studies BMI as continuous outcome showed that higher BMI was associated with composite poor outcome with BMI mean difference between groups 0.55 kg/m^2^ (95% CI 0.07–1.03, P=0.02) with I^2^=92.65%. Meta-analysis from 3 studies with BMI as dichotomous outcome showed that BMI ≥30 kg/m^2^ (obese) was associated with composite poor outcome with OR 1.89 (95% CI 1.03−3.34, P=0.03) with I^2^=-13.2%. Subgroup analysis was not performed due to the limitation number of studies for several outcomes. The results of meta-analysis are summarized in Figure 2.

**Figure 2.**
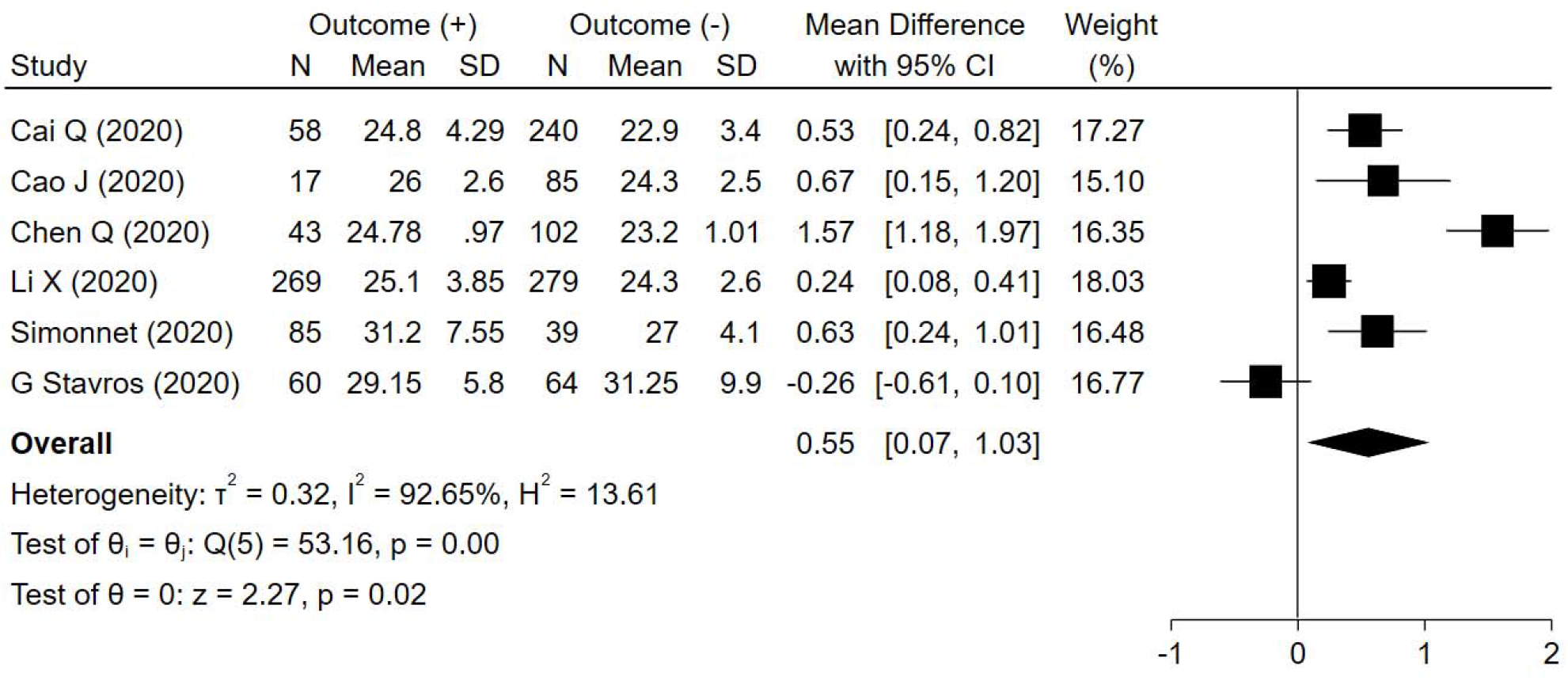
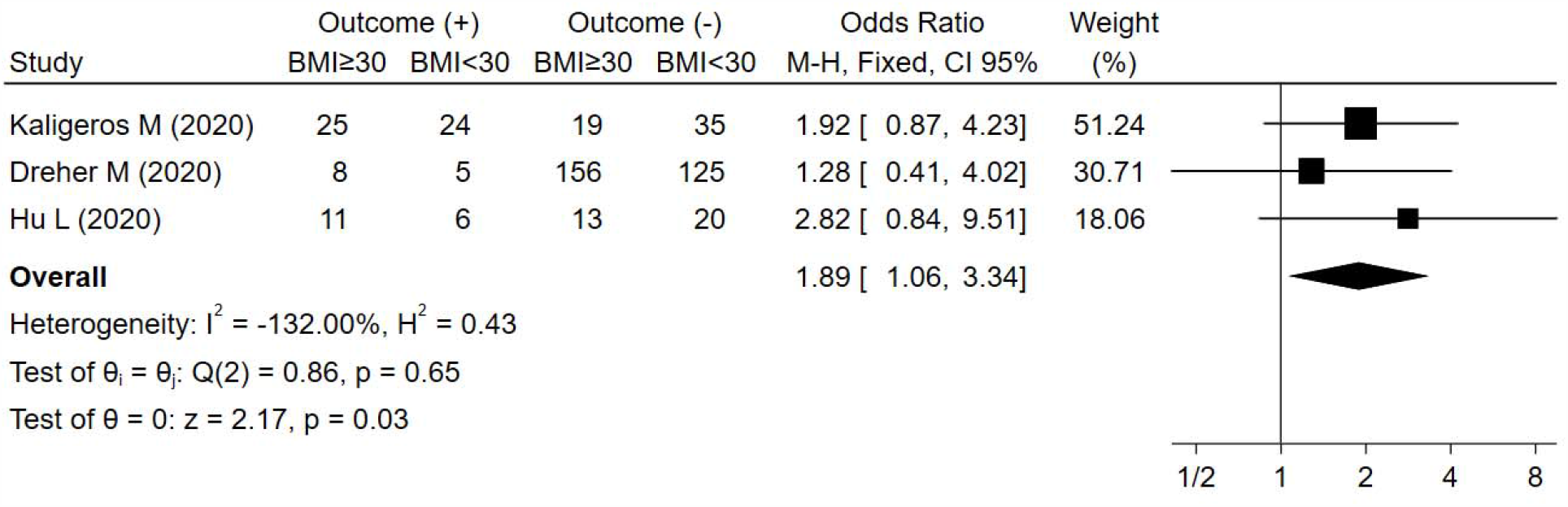
Obesity and Composite Poor Outcome. Forest plot presented with BMI both as continuous outcome and dichotomous outcome. BMI cut-off for dichotomous outcome defined as ≥30 kg/m^2^. Subgroup analysis was not performed due to limitation number of studies. Forest plot shows obesity was associated with increased composite poor outcome.

### 3.4 Meta-Regression

Meta-regression from BMI as continuous outcome in Figure 3 showed the association between higher BMI and composite poor outcome was affected by age with regression coefficient =-0.06 and P=0.02. Hypertension and DM type 2 were not associated with composite poor outcome with P=0.057 and p=0.101. Meta-regression from BMI as dichotomous (obese vs non-obese) outcome in Figure 4 outcome that hypertension, DM type 2, and age were not associated with composite poor outcome with P=0.412, P=0.354, and P=0.512 respectively.

**Figure 3.**
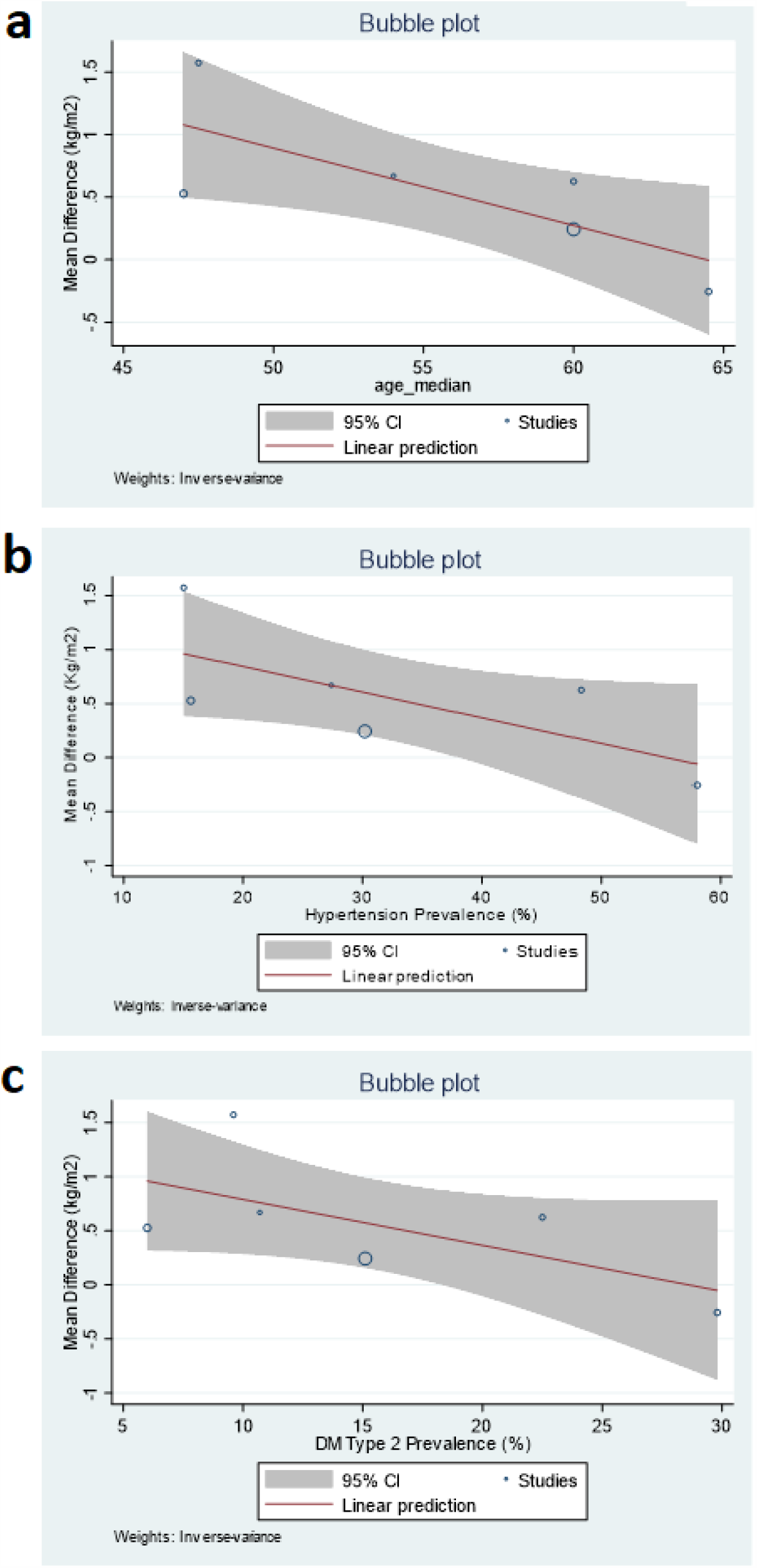
Bubble-Plot for meta-regression for BMI presented as continuous outcome. Meta-regression analysis showed that the association between obesity and composite poor outcime was affected by age [A] with p=0.02

**Figure 4.**
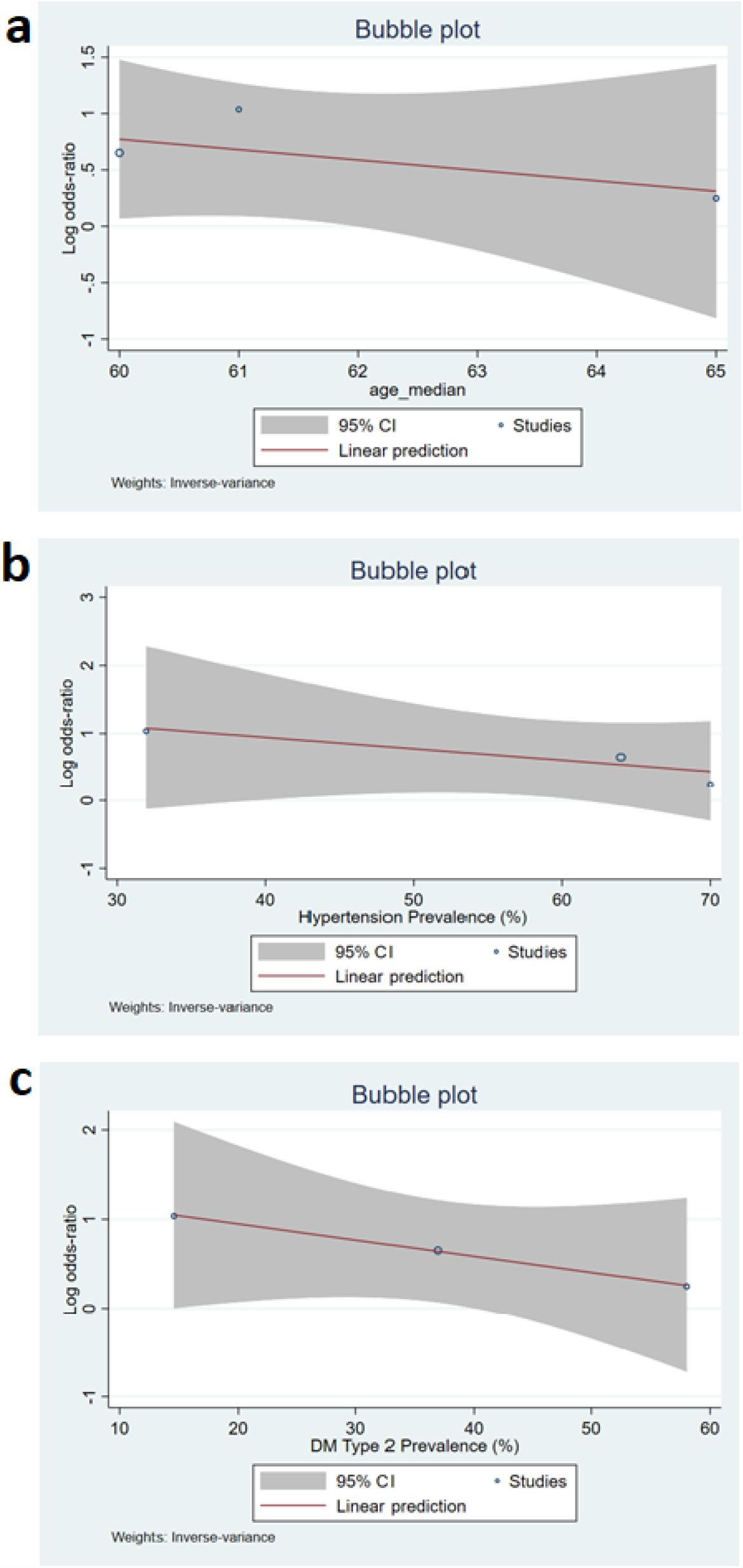
Bubble-Plot for meta-regression for BMI presented as dichotomous outcome. Meta-regression analysis showed that the association between obesity and composite poor outcome was not affected by age, hypertension, and DM type 2.

### 3.5 Publication Bias

The funnel plot graph in Figure 5 for BMI as continue outcome showed asymmetrical non-inverted funnel that may indicate the presence of publication bias. Thus, a more formal evaluation of funnel plot asymmetry using Egger test for continuous outcome was conducted. Egger test showed P=0.374 indicates that there is no evidence of small-study effects.

**Figure 5.**
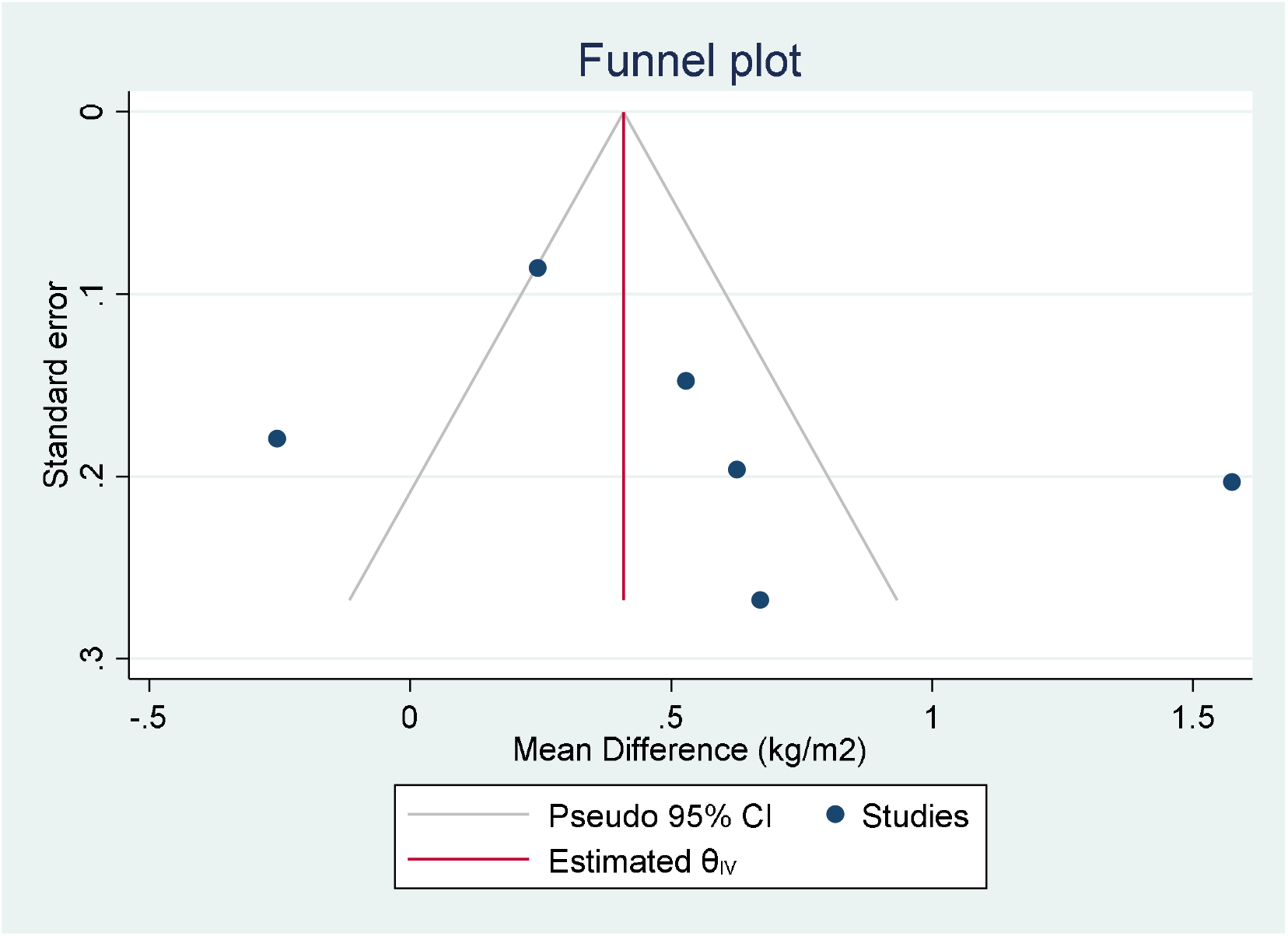
Funnel Plot for BMI as continuous outcome. Funnel plot shows asymetrical non-inverted funnel. Egger tes was conducted with p=0.374

## 4. Discussion

Body Mass Index (BMI) is used to classify obesity. BMI is calculated as the ratio of weight in kilograms to the square of height in meters, expressed in units of kg/m2. According to WHO, BMI was classified into six groups: underweight (<18.5 kg/m^2^), normal (18.5 – 24.9 kg/m^2^), pre-obesity (25 – 29.9 kg/m^2^), obesity class I (30 – 34.9kg/m^2^), obesity class II (35 – 39.9kg/m^2^), and obesity class III (>40 kg/m^2^).^18^. The worldwide prevalence of overweight (BMI ≥ 25 kg/m^2^) and obesity has increased significantly and extent that nearly a third of the world population. The trend in obesity has increased in both and adults and children of all ages with The American and The European were two regions with the highest prevalence of obesity.^19, 20^. Although China has similar incidence compared to US. ^21^ Obesity can be seen as a pandemic. According to Centers for Disease Control and Prevention (CDC), the age-adjusted prevalence of obesity was 42.4% and 9.4% for severe obesity.^22^

This meta-analysis study showed that in COVID-19 patients with composite poor outcome which consist of mortality, morbidity, admission of ICU, mechanical ventilation, ARDS, and severe Covid-19 had higher BMI compared to patients without composite poor outcome patients. Meta-regression showed that this association was influenced by age. This association weaker in older patients. Obesity inreased the risk for composite poor outcome with OR 1.89. Previous data showed that obesity was present in nearly one-third of hospitalizations and fatal cases during the 2009 H1N1 pandemic and recognized as an independent risk factor for severity, hospitalization, increased risk of transmission, and mortality of influenza during the H1N1 pandemic in 2009. ^23, 24^ Therefore, is not surprising that higher BMI and obesity as is also associated with poor outcome in SARS-CoV-2 infection as well.

Obesity has been identified as a significant risk factor for severe disease following lower respiratory tract infections. The mechanical effects of obesity can cause airway narrowing and increased resistance. In addition, airway narrowing in obesity correlates with airway closure and airway hyperresponsiveness (AHR)^5^ Obesity appears to be associated with a higher chance to developing respiratory complication and need for ICU admission and mechanical ventilation.^25^ COVID-19 can lead to potential airways threatening and result in ARDS and respiratory failure.^24^ Respiratory muscle strength decreases and oxygen demand increase more than three-fold due to increased airway resistance and chest wall mechanics.^26^ Increased oxygen consumption can lead to respiratory failure and predispose the need for more oxygen support. In addition, due to anatomical factor obese patients have a higher incidence difficult mask ventilation compared to non-obese patients. ^27^

Other factor that probably explains the association between obesity and poor outcome of COVID-19 is that obesity has a negative effect on the immune system and host defense mechanism.^28^ Adipose tissue involved in many physiologic and metabolic processes as well as a reservoir for T lymphocytes and macrophages. Excess body fat reduces the response to antiviral agents through poor T cell and macrophage functions.^29^ In other hand, obesity is also associated with chronic activation of the innate immune system related to local and systemic inflammation^28^ Adipose tissues secrete adipokines which differentially expressed between obese and lean subject. Patients who are obese experience chronic alterations in circulating inflammatory mediators derived from adipokines.^30^ Other inflammatory mediators that are increased in obesity include tumor necrosis factor alpha (TNF-α), interleukin (IL-) 8 and IL-6, high-sensitivity C-reactive protein (hs-CRP) and monocyte chemoattractant protein-1 (MCP-1).^6^ Increased inflammatory cytokines leading to cytokine storm was also reported in some studies. ^31^ As proposed by Siddiqi, a minority of COVID-19 patients will transition into the third and most severe stage of illness, which manifests as an extra-pulmonary systemic hyper inflammation syndrome. In this stage, markers of systemic inflammation appear to be elevated. COVID-19 infection results in a decrease in helper, suppressor and regulatory T cell counts. Inflammatory cytokines and biomarkers such as interleukin (IL)-2, IL-6, IL-7,granulocyte-colony stimulating factor, macrophage inflammatory protein 1-α, tumor necrosis factor-α, C reactive protein, ferritin, and D-dimer are significantly elevated in those patients with more severe disease.^32^ It can be assumed that obese COVID-19 patients more likely to fall to this stage

At present, there is no definitive therapy for COVID-19 treatment. Prevention of disease transmission remain most important concern. The fact that prevalence of overweight and obesity extent nearly a third of the world population, seems like we have to face two pandemics simultaneously. A healthy lifestyle, particularly achieving ideal body weight, is very important in preventing and reducing composite poor outcome including mortality if a person contracted to COVID-19. Stakeholders have an obligation to encourage the community to implement a healthy lifestyle to reduce the prevalence of overweight and obesity especially during COVID-19 pandemic.

## 5. Conclusion

Obesity is a risk factor of composite poor outcome of COVID-19. On the other hand, COVID-19 patients with composite poor outcome have higher BMI. BMI should always be assessed in every COVID-19 patient and special attention should be given in the management of obese patients.

## Data Availability

The data used to support the findings of this study are included within the article

## Acknowledgment

AYS, NNS, and P conceived and designed the study. PS, HS, IDK, FF acquire the data. AYS and P performed data extraction and interpreted the data. Ays and P performed the statistical analysis. All authors contributed to the writing of the manuscript.

## References

1. WHO. Coronavirus disease 2019 (COVID-19) Situation Report – 127. 2020.

2. Kruglikov IL, Scherer PE. The role of adipocytes and adipocyte-like cells in the severity of COVID-19 infections. Obesity. n/a (n/a):

3. Ge H, Wang X, Yuan X, Xiao G, Wang C, Deng T, et al. The epidemiology and clinical information about COVID-19. Eur J Clin Microbiol Infect Dis. 2020; 1-9

4. Guo Y-R, Cao Q-D, Hong Z-S, Tan Y-Y, Chen S-D, Jin H-J, et al. The origin, transmission and clinical therapies on coronavirus disease 2019 (COVID-19) outbreak - an update on the status. Mil Med Res. 2020; 7 (1): 11-

5. Kulcsar KA, Coleman CM, Beck SE, Frieman MB. Comorbid diabetes results in immune dysregulation and enhanced disease severity following MERS-CoV infection. JCI Insight. 2019; 4 (20):

6. Dixon AE, Peters U. The effect of obesity on lung function. Expert Review of Respiratory Medicine. 2018; 12 (9): 755–67

7. Wan X, Wang W, Liu J, Tong T. Estimating the sample mean and standard deviation from the sample size, median, range and/or interquartile range. BMC Medical Research Methodology. 2014; 14 (1): 135

8. Higgins J, Wells G. Cochrane handbook for systematic reviews of interventions. Edisi Vol Wiley Online Library; 2011.

9. Cai Q, Huang D, Ou P, Yu H, Zhu Z, Xia Z, et al. COVID-19 in a designated infectious diseases hospital outside Hubei Province, China. Allergy. 2020;

10. Cao J, Tu WJ, Cheng W, Yu L, Liu YK, Hu X, et al. Clinical Features and Short-term Outcomes of 102 Patients with Corona Virus Disease 2019 in Wuhan, China. Clinical infectious diseases: an official publication of the Infectious Diseases Society of America. 2020;

11. Chen Q, Zheng Z, Zhang C, Zhang X, Wu H, Wang J, et al. Clinical characteristics of 145 patients with corona virus disease 2019 (COVID-19) in Taizhou, Zhejiang, China. Infection. 2020;

12. Li X, Xu S, Yu M, Wang K, Tao Y, Zhou Y, et al. Risk factors for severity and mortality in adult COVID-19 inpatients in Wuhan. The Journal of allergy and clinical immunology. 2020;

13. Simonnet A, Chetboun M, Poissy J, Raverdy V, Noulette J, Duhamel A, et al. High prevalence of obesity in severe acute respiratory syndrome coronavirus-2 (SARS-CoV-2) requiring invasive mechanical ventilation. Obesity (Silver Spring, Md). 2020;

14. Kalligeros M, Shehadeh F, Mylona EK, Benitez G, Beckwith CG, Chan PA, et al. Association of Obesity with Disease Severity among Patients with COVID-19. Obesity (Silver Spring, Md). 2020;

15. Hu L, Chen S, Fu Y, Gao Z, Long H, Wang JM, et al. Risk Factors Associated with Clinical Outcomes in 323 COVID-19 Hospitalized Patients in Wuhan, China. Clinical infectious diseases: an official publication of the Infectious Diseases Society of America. 2020;

16. Dreher M, Kersten A, Bickenbach J, Balfanz P, Hartmann B, Cornelissen C, et al. The Characteristics of 50 Hospitalized COVID-19 Patients With and Without ARDS. Dtsch Arztebl International. 2020; 117 (16): 271–8

17. Memtsoudis SG, Ivascu NS, Pryor KO, Goldstein PA. Obesity as a risk factor for poor outcome in COVID-19 induced lung injury: the potential role of undiagnosed obstructive sleep apnoea. British journal of anaesthesia.

18. WHO. 2020 [Tersedia dari: http://www.euro.who.int/en/health-topics/disease-prevention/nutrition/a-healthy-lifestyle/body-mass-index-bmi.

19. Chooi YC, Ding C, Magkos F. The epidemiology of obesity. Metabolism: clinical and experimental. 2019; 92 6-10

20. Michalakis K, Goulis DG, Vazaiou A, Mintziori G, Polymeris A, Abrahamian-Michalakis A. Obesity in the ageing man. Metabolism: clinical and experimental. 2013; 62 (10): 1341–9

21. Hu C, Jia W. Diabetes in China: Epidemiology and Genetic Risk Factors and Their Clinical Utility in Personalized Medication. Diabetes. 2018; 67 (1): 3–11

22. Prevalence of Obesity and Severe Obesity Among Adults: United States, 2017–2018. Center for Disease Control and Prevention, 2020.

23. Moser JS, Galindo-Fraga A, Ortiz-Hernandez AA, Gu W, Hunsberger S, Galan-Herrera JF, et al. Underweight, overweight, and obesity as independent risk factors for hospitalization in adults and children from influenza and other respiratory viruses. Influenza and other respiratory viruses. 2019; 13 (1): 3–9

24. Honce R, Schultz-Cherry S. Impact of Obesity on Influenza A Virus Pathogenesis, Immune Response, and Evolution. Frontiers in immunology. 2019; 10 1071

25. Severin R, Arena R, Lavie CJ, Bond S, Phillips SA. Respiratory Muscle Performance Screening for Infectious Disease Management Following COVID-19: A Highly Pressurized Situation. The American journal of medicine. 2020;

26. Sood A. Altered resting and exercise respiratory physiology in obesity. Clinics in chest medicine. 2009; 30 (3): 445-54, vii

27. Moon TS, Fox PE, Somasundaram A, Minhajuddin A, Gonzales MX, Pak TJ, et al. The influence of morbid obesity on difficult intubation and difficult mask ventilation. Journal of anesthesia. 2019; 33 (1): 96–102

28. Frasca D, McElhaney J. Influence of Obesity on Pneumococcus Infection Risk in the Elderly. Frontiers in endocrinology. 2019; 10 71

29. Coelho M, Oliveira T, Fernandes R. Biochemistry of adipose tissue: an endocrine organ. Archives of medical science: AMS. 2013; 9 (2): 191–200

30. Hibbert K, Rice M, Malhotra A. Obesity and ARDS. Chest. 2012; 142 (3): 785–90

31. Zhang X, Wu K, Wang D, Yue X, Song D, Zhu Y, et al. Nucleocapsid protein of SARS-CoV activates interleukin-6 expression through cellular transcription factor NF-κB. Virology. 2007; 365 (2): 324–35

32. Siddiqi HK, Mehra MR. COVID-19 illness in native and immunosuppressed states: A clinical-therapeutic staging proposal. The Journal of heart and lung transplantation: the official publication of the International Society for Heart Transplantation. 2020; 39 (5): 405–7

